# Knowledge about benefits and risks of undergoing cataract surgery among cataract patients in Southern China

**DOI:** 10.1101/19014100

**Authors:** Guofang Ye, Bo Qu, Wen Shi, Xin Chen, Pengjuan Ma, Yuxin Zhong, Shida Chen, Ecosse Lamoureux, Yingfeng Zheng

**Author notes:** **Address for correspondence:** Yingfeng Zheng, MD, PhD, Zhongshan Ophthalmic Center, Guangzhou, People’s Republic of China. **Financial support:** This study was funded by the National Natural Science Foundation of China (81530028; 81721003), Local Innovative and Research Teams Project of Guangdong Pearl River Talents Program; the State Key Laboratory of Ophthalmology, Zhongshan Ophthalmic Center, Sun Yat-sen University. These authors contributed equally to this work (Guofang Ye, Bo Qu).

## Abstract

**Objective:** To develop a theoretical framework for assessing knowledge about the possible outcomes of undergoing cataract surgery, and explore the association of knowledge level with psychological status and decision quality among patients with cataract in Southern China.

**Methods:** The details of the knowledge scale were based on the health education information booklet provided by National Eye Institute, NIH. We used a theory-based approach to assess gist knowledge, which comprises conceptual and numeric questions related to knowledge of the possible surgical outcomes. The scale was then used in a cross-sectional study to assess the association of knowledge score with psychological status and decision quality of cataract patients, including worry, anxiety, attitudes, intentions, decisional conflict, confidence in decision making, anticipated regret and temporal orientation.

**Results:** A total of 489 participants with age-related cataract were included in this study, and 10.2% (50/489) of them had adequate level of knowledge. The knowledge scale was significantly associated to the levels of worry (Odds Ratio (OR) = 0.36, 95%CI: 0.18, 0.70; *P* = 0.003), anxiety (beta coefficient = -5.36, 95%CI: -8.88,-1.84; *P* = 0.003), inaction regret (OR = 0.49, 95%CI : 0.28, 0.88; *P* = 0.016) and decision conflict (beta coefficient = -7.93, 95%CI: -12.81, -3.04; *P* = 0.002) in multivariate analyses adjusted for age, sex, education level and literacy level.

**Conclusion:** The level of knowledge adequacy with cataract surgery outcomes is high in China and was associated with psychological status and decision quality. These findings suggest that strategies targeting knowledge of possible surgical outcomes may reduce psychological stress and improve decision quality among patients with age-related cataract.

## 1. Introduction

Cataract is the leading cause of visual impairment (VI) and blindness.^1^ Surgical removal of the clouded lens remains one of the most commonly used procedures worldwide^2 3^. Modern cataract surgery (e.g., phacoemulsification) has become a safe and reliable therapeutic procedure, and there is an increasing number of people with mild VI or even good vision receiving the surgery for the improvement of their vison and quality of life^4^.Therefore, it is essential to assess patients’ knowledge of the possible outcomes of undergoing or delaying cataract surgery, which might inform the clinical option that best accommodates their personal preferences and making informed choices^5^. Although knowledge is one of the most commonly used outcome measures in decision aid trials^6^, none of the currently available tools adequately captures the level of knowledge and determine the effect of a decision supporting program on the quality of decision for cataract patients.^7-9^

Furthermore, little is known about the association between cataract patients’ knowledge; and their psychological status and decision quality. This is a clinically relevant issue, as mental health problems have become a huge social burden among the elderly population.^10^ Recent evidence suggests that cataract-related vision loss has a great impact on patients’ psychological parameters,^11 12^ and patients with age-related cataract show a higher rate of depression and anxiety before surgery^13^. All these adverse psychological factors among the elderly may contribute to worse physical health status, and even high suicide risk.^14^ Given that knowledge is a potentially modifiable factor, improving cataract knowledge for elderly people may have broad strategic implications for alleviating their mental health problems before surgery.

Finally, with the increasing demand for shared decision-making in clinical guidelines, there is a great emphasis on increasing patients’ knowledge which can lead to improved decision quality.^15 16^ A systematic review containing over 100 randomized controlled trials worldwide suggested that patient decision aids increase knowledge and accurate risk perceptions; and reduce decisional conflict and indecision about personal values, but it has divergent results on decision confidence and anticipated regret.^8^ Considering the data vary on the patients’ knowledge and their decision quality in different countries, it is necessary to investigate this potential link among cataract patients.

The primary purpose of the current study was to develop a theoretical framework for assessing the magnitude of knowledge associated with cataract surgery decision in China. In addition, we explored the potential association of patients’ knowledge with their psychological status (including cataract worry and anxiety), attitudes and intentions towards surgery and decision quality (including anticipated regret, decisional conflict, confidence in decision making and temporal orientation).

## 2. Method

### 2.1 Development and design of a knowledge scale for decisions about the timing of cataract surgery

We determined the content of the knowledge scale using an expert-led approach based on the health education information booklet provided by National Eye Institute, NIH.^17^

The health information booklet mentions that:

“A cataract needs to be removed only when vision loss interferes with your everyday activities, such as driving, reading, or watching TV. You and your eye care professional can make this decision together. Once you understand the benefits and risks of surgery, you can make an informed decision about whether cataract surgery is right for you. In most cases, delaying cataract surgery will not cause long-term damage to your eye or make the surgery more difficult. You do not have to rush into surgery.” ^17^

These statements highlight two important information relevant to surgical decision that should be revealed to patients: the benefits and risks of surgery. We therefore developed key knowledge items based on Fuzzy Trace Theory^18 19^, which proposes two ways in which people process information: (1) processing and remembering the underlying meaning (“gist”) and (2) processing and remembering precise details (“verbatim”). We argued that when people are making decisions, gist information plays a more important role than verbatim information, because the former relies less on accurate details. We therefore proposed 10 knowledge items to reflect whether patients understood the gist of the information on conceptual knowledge of the definition of cataract, as well as the possible impacts of undergoing or delaying cataract surgery. Since numeracy may contribute to poor understanding of health information and influence the extent of medical decision making^20^, we also included two numeric questions to assess the potential number of people affected by undergoing or delaying cataract surgery (Table 1).

**Table 1.**
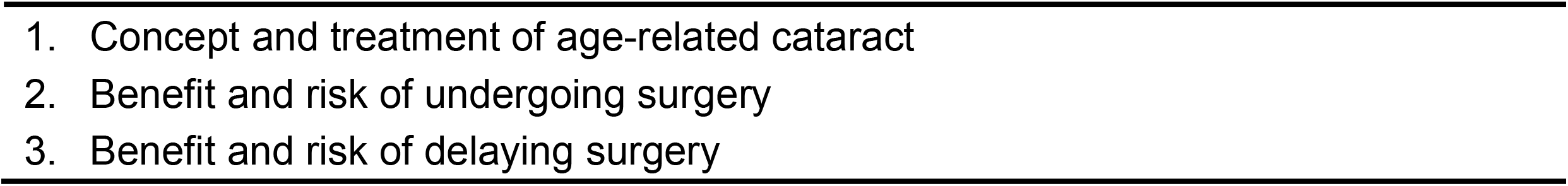
Core knowledge items.

Marks for all questions were summed to give a maximum total score of 14 (Table 2). We proposed an *a priori* definition of “adequate knowledge” when responders scored at least 50% of available marks on subscale 1; and 40% on subscales 2 and 3, including at least one numerical mark.

**Table 2.** Marking scheme for cataract surgery knowledge questionnaire.

| Core knowledge items | Item | Conceptual and numeric knowledge questions | Actual answers | Marking/scoring scheme | Maximum mark awarded |
| --- | --- | --- | --- | --- | --- |
| Subscale 1: Questions about cataract and cataract surgery | Q1 | In general, age-related cataracts tend to “grow” quickly, so vision gets worse rapidly. | (1) Yes<br>(2) No | (2) = 1 mark | 4 |
|  | Q7 | Who is the suitable person to make decision about how to treat age-related cataract? | (1) You<br>(2) You and your doctor can make the decision together<br>(3) Your doctor | (2) =1 mark |  |
|  | Q9 | What is age-related cataract? | (1) As we age, some of the protein may clump together and start to cloud a small area of the lens, making it harder to see.<br>(2) As we age, we feel that some mosquitoes flying in our eyes. | (1) =1 mark |  |
|  | Q10 | Some medication or eyedrops are available for the treatment of age-related cataract. | (1) Yes<br>(2) No | (2) =1 mark |  |
| Subscale 2: Questions about undergoing surgery | Q2 | In general, ALL cataract patients who have cataract surgery have better vision afterward. | (1) Yes<br>(2) No | (2) = 1 mark | 5 |
|  | Q5 | When is the right time to have cataract surgery? | (1) When vision loss due to cataract has already affected your daily activities.<br>(2) When vision loss due to cataract has not yet affected your daily activities. | (1) =1 mark |  |
|  | Q8 | Cataract surgery significantly increases your risk of retinal detachment and blindness after cataract surgery. | (1) Yes<br>(2) No | (2) = 1 mark |  |
|  | Q11 | Number of cataract patients out of 100 whose vision will be significantly improved, if they undergo cataract surgery? | The participant is asked to give the exact number | 80-95 = 2 marks<br>51-79 = 1 mark<br>95-99 = 1 mark<br>0-50 = 0 mark<br>100 = 0 mark |  |
| Subscale 3: Questions about delaying surgery | Q3 | In general, delaying cataract surgery will cause long-term damage and you will end up with blindness | (1) Yes<br>(2) No | (2) = 1 mark | 5 |
|  | Q4 | In general, delaying cataract surgery will make it harder for your surgeon to perform cataract surgery. | (1) Yes<br>(2) No | (2) = 1 mark |  |
|  | Q6 | If you choose delaying cataract surgery now, it is important to receive follow-up monitoring. | (1) Yes<br>(2) No | (1) = 1 mark |  |
|  | Q12 | Number of cataract patients out of 100 whose vision does not deteriorate significantly over the next 5 years, if they delay cataract surgery? | The participant is asked to give the exact number | 20-80 = 2 marks<br>81-99 = 1 mark<br>1-19 = 1 mark<br>0 = 0 mark<br>100 = 0 mark |  |

### 2.2 Implementing the knowledge scale in a community-based eye disease screening setting

We next applied the knowledge scale in a community-based and cross-sectional study conducted at Yuexiu district of Guangzhou, China, between June 12 and July 23 in 2019. We recruited participants via telephone survey in Huanghuagang Block,^21^ a socio-economically representative area in urban Southern China. The study adhered to the Declaration of Helsinki and approvals were obtained from the Zhongshan Ophthalmic Center Institutional Review Board, and written informed consent was obtained from the participants.

#### 2.2.1 Inclusion and exclusion criteria

All persons aged 50 to 80 years, presenting to the eye disease screening site were eligible for study participation if they had been definitely diagnosed with an age-related cataract but not received previous cataract surgery, and they were willing to participate in this study and provide written informed consent. We excluded subjects if they were bilaterally blind (presenting distance visual acuity worse than 3/60), suffered from ocular comorbidities which could not be cured by cataract surgery alone, had contraindications of cataract surgery, or had self -reported mental disorders or hearing impairment that affect face-to-face communication with interviewers.

#### 2.2.2 Data collection

We used a detailed interviewer-administered questionnaire to collect participants’ information on demographics, knowledge about cataract and other variables, including cataract worry and anxiety, attitudes towards cataract surgery^22^, intentions^23^, decisional conflict, confidence in decision making, anticipated regret (action regret and inaction regret) and temporal orientation^9 24^.

All participants involved in our study were required to complete the questionnaires under the guidance of trained interviewers from an independent non-profit company. To enhance the reliability of the answers provided in this survey, our interviewers, who have been trained to make personal connections with elderly peoples and answer their questions any time, explained the questions aloud one by one to make sure the information provided was well-understood by every subject, including those with low level of literacy and education.

#### 2.2.3 Knowledge assessment

Knowledge was assessed as a dichotomous outcome based on the marking scheme (Table 2).

#### 2.2.4 Assessment of psychological status and decision quality

##### Psychological status

Patients’ anxiety was measured with the six-item Spielberger State Trait Anxiety Inventory (STAI),^25 26^ one of the most commonly used measures of anxiety in research and clinics. The version used here was translated into Chinese and its validity has been demonstrated in many studies.^27^All items were rated on a 4-point scale ranging from “almost never” to “almost always” (scored 1-4). A total score ranged from 4 to 24 with higher scores indicating greater anxiety^28^. Furthermore, participants were asked how worried they were of vision impairment due to cataract; they responded on a 4-point Likert scale anchored from “not worried at all” to “very worried” (scored 1 to 4 respectively).^9 29^.

##### Attitudes and intentions

We measured patients’ attitudes towards cataract surgery with a validated, theory-based generic attitudes scale comprising six items with five response categories (ranging from “strongly disagree” to “strongly agree”), forming a scale from six to 30^24^. As reported previously, we set the threshold for a positive attitude at 24^9 24^. Patients indicated their intentions about receiving cataract surgery as soon as possible via a single item with five responses ranging from “definitely will not” to “definitely will” (scored 1-5)^9 24^. In our survey, we classified “definitely will” and “likely to” receive surgical treatment as positive intentions.

##### Decisional conflict

We used a scale adapted from the Ottawa Decisional Conflict Scale (DCS) to measure patients’ overall decisional conflict, the particular factors causing uncertainty (e.g. feeling uninformed, unclear about personal values and unsupported in decision making) and perceived effective decision making (16 items, 5-point scale ranging from 0 to 4).^30^ The reliability and validity of the Chinese version of the DCS were demonstrated previously.^31^

##### Decision confidence

We used the Decision Self-Efficacy Scale (DSES) to assess patients’ confidence in obtaining adequate information during decision making process. The scale for patients has demonstrated good internal consistency with a Cronbach’s α of 0.84.^32^. Participants were asked how confident they felt when making an informed choice on a 20-item scale with 5 response categories ranging from “not at all confident” to “very confident” (scored 1-5). The total scores ranged from 0 to 100, and higher scores indicated greater self-efficacy^9^.

##### Anticipated regret

We used two items adapted from the scale in previous studies to assess participants’ anticipated regret about receiving cataract surgery (action regret) and about delaying cataract surgery (inaction regret), with five response categories ranging from “strongly disagree” to “strongly agree” (scored 1-5)^9 33^. In this study, we assessed the level of inaction regret and action regret as “no” (1-2 scores), “unsure” (3 scores) and “yes” (4-5 scores).

##### Temporal orientation

Temporal orientation, defined by the tendency to emphasize psychological past, present and future, affects patients’ emotion and decision making process.^34^ We used the four-item short form of the Consideration of Future Consequences Scale to measure this variable of participants^35^, with five response categories ranging from “strongly disagree” to “strongly agree” (scored 1 - 5). The total scores ranged from 5 to 25, with higher scores indicating a long-term temporal orientation (i.e., greater orientation towards the future)^9 36^.

### 2.3 Statistical analyses

Data analyses were performed using Stata version 12.0. The associations between adequate knowledge and other demographic variables (age, sex, education level, literacy, BCVA and insurance type) were examined using Chi-square or Fisher’s exact tests. A potential multicollinearity between education level and literacy was assessed using variance inflation factor (VIF), which is less than 1.5 in the regression analysis. For categorical variables, we developed univariate logistic regression to calculate the odds ratio (OR) and the 95% confidence interval (CI) for the associations between knowledge level and other factors (e.g., attitudes, intentions, worry and anticipated regret) adjusted for age, sex, education level and literacy. For continuous variables, we developed simple linear regression to examine the associations between patients’ knowledge level and other factors (e.g., anxiety, decision conflict, decision confidence and temporal orientation). All P-values were 2-sided and considered statistically significant if less than 0.05.

## 3 Results

A total of 489 subjects participated in our study. The mean (± standard Deviation) age of our participants was 64.06±5.46 years (Table 3). Most participants were aged 60∼70 years (65.24%), female (70.14%), had higher school certificate or above (71.99%), semiliterate or literate (92.23%), had the best corrected visual acuity (BCVA) of the worse-seeing eye greater than logMAR 20/40 (89.78%), and had public medical insurance (98.98%). There were no significant differences for the participants’ sociodemographic characteristics such as age, gender, education level, literacy, BCVA, and insurance between participants who had adequate and inadequate knowledge (all with P > 0.05). Overall, 439 (89.78%) of our study participants had inadequate knowledge about cataract surgery and 50 (10.22%) had adequate knowledge.

**Table 3.**
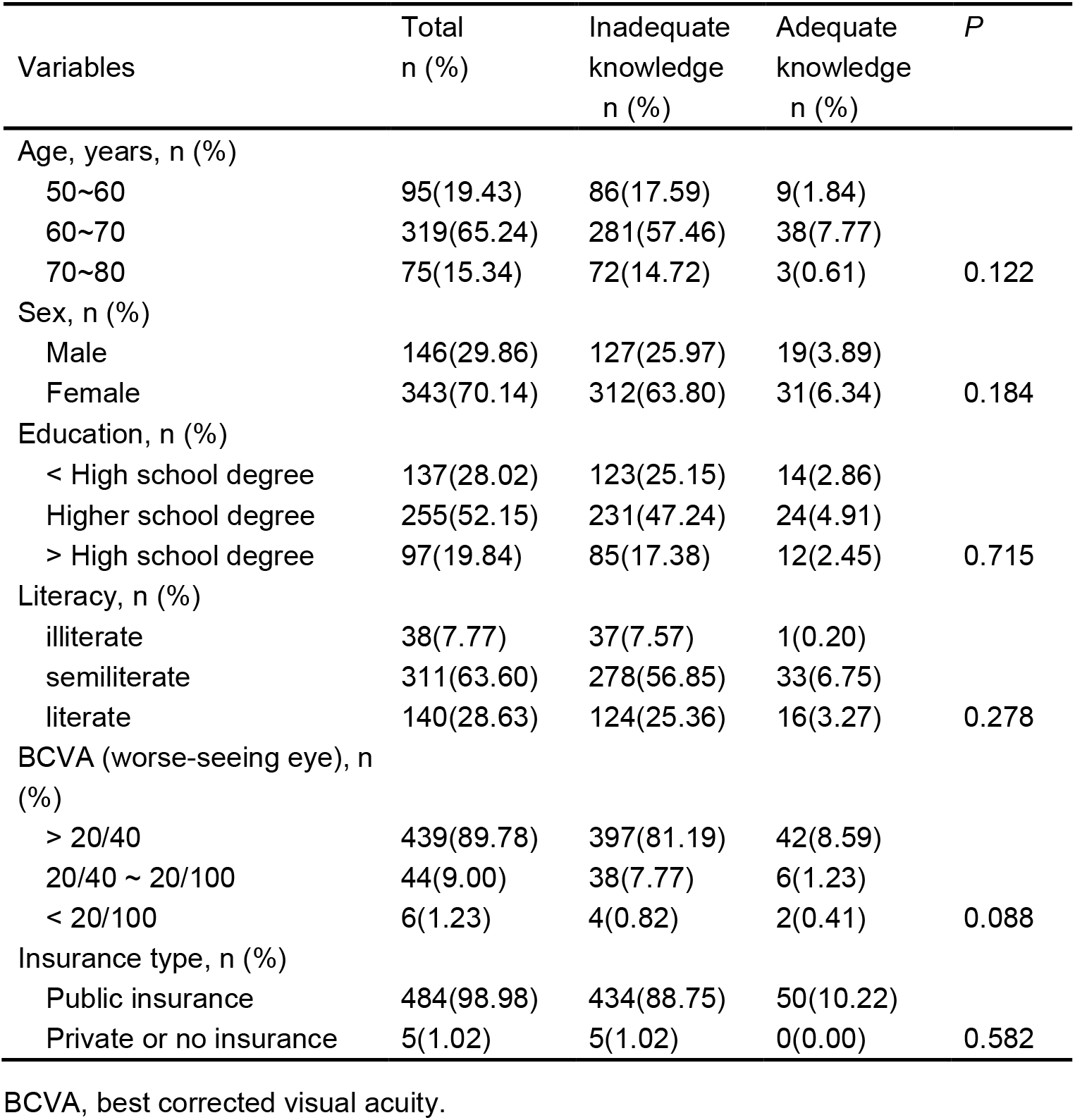
Sociodemographic characteristics of participants (n=489) in our survey.

In Tables 4 - 6, each value represents the result of a separate regression model, with attitudes, intentions, cataract worry, anxiety, anticipated regret, decisional conflict, confidence and temporal orientation as the dependent variable, and age, sex, education level and literacy level as co-variates in the multivariate regression models. There was no statistically significant interaction between education level and literacy when we included both of them in the same multivariate model (*P* > 0.05, data not shown).

**Table 4.** Association of knowledge with patients’ attitudes and intentions.

|  | Attitudes (ref = positive) |  | Intentions (ref = not receiving surgery) |  |
| --- | --- | --- | --- | --- |
|  | OR (95% CI)* | *P* value | OR (95% CI)* | *P* value |
| Knowledge | 4.85 (2.10, 11.25) | < 0.001 | 0.30 (0.13, 0.73) | 0.008 |

OR, odds ratio; 95% CI, confidence interval.
\* Multivariate analysis adjusted for age, sex, education level, and literacy level.

**Table 5.** Association of knowledge with patients’ psychological status.

|  | Worry (ref = no) |  | Anxiety |  |
| --- | --- | --- | --- | --- |
|  | OR (95% CI)* | *P* value | Regression coefficient (95% CI)* | *P* value |
| knowledge | 0.36 (0.18, 0.70) | 0.003 | -5.36 (-8.88, -1.84) | 0.003 |

OR, odds ratio; 95% CI, confidence interval.
\* Multivariate analysis adjusted for age, sex, education level, and literacy level.

**Table 6.** Association of knowledge with patient’ decision quality.

|  | Inaction Regret (ref = no) |  | Action regret (ref = no) |  | Decisional conflict |  | Confidence |  | Temporal orientation |  |
| --- | --- | --- | --- | --- | --- | --- | --- | --- | --- | --- |
|  | OR (95%CI)* | *P*<br>value | OR (95%CI)* | *P*<br>value | Regression<br>coefficient (95%CI)* | *P*<br>value | Regression<br>coefficient (95%CI)* | *P*<br>value | Regression<br>coefficient (95%CI)* | *P*<br>value |
| knowledge | 0.49 (0.28, 0.88) | 0.016 | 1.45 (0.81, 2.56) | 0.211 | -7.93 (-12.81, -3.04) | 0.002 | -0.28 (-4.63, 4.07) | 0.899 | 0.15 (-0.45, 0.75) | 0.624 |

OR, odds ratio; 95% CI, confidence interval.
\* Multivariate analysis adjusted for age, sex, education level, and literacy level.

Table 4 shows the correlations of knowledge with attitudes and intentions of treatment options. Adequate level of knowledge was found to correlate with negative attitudes (*P* < 0.001) and the intention to delay cataract surgery (*P* = 0.008). The associations of knowledge with cataract worry and anxiety are shown in Table 5. In our study, we found that participants with adequate level of knowledge were less likely to have cataract worry and anxiety (*P* = 0.003 for both). In terms of decision quality shown in Table 6, adequate level of knowledge was negatively associated with decision conflict (*P* = 0.002) and inaction regret (*P* = 0.016). Other factors about decision quality including action regret, decision confidence and temporal orientation were not associated with patients’ knowledge (all with *P* > 0.05).

## 4 Discussion

This study describes the development of a knowledge scale for measuring the benefits and risks of undergoing or delaying cataract surgery. The measure consisted of three core knowledge items considered indispensable for making an informed choice about cataract surgery. Our study provided evidence that the knowledge scale was responsive for assessing gist knowledge of cataract surgery. In addition, we suggested that cataract patients with adequate knowledge were more likely to possess greater psychological conditions and decisional quality, although they tended to be more negative and delay surgical treatment.

Our knowledge scale was an objective measurement developed under the guidance of experts using a number of conceptual and numeric questions rather than a subjective, self-reported assessment of knowledge, because the latter might result in many false positives.^37^ Given that public awareness of the benefits and risks of cataract surgery is limited, we adopted the expert-led approach in the current study. However, it contrasts with other studies that both patients and professionals were involved in the establishment of the scale, which, to some extent, could address patients’ concerns and minimize their misunderstanding of the sentences.^38 39^ To address these questions, we chose not to use the confusing or difficult words in the questionnaire and referred to content domains for knowledge measurement in previous study.^38^ Furthermore, we set a cut-off for adequate knowledge (i.e. 50% or above) using a competency-based method,^38 40^ enabling us to assess knowledge based on pre-determined standards rather than participants’ ranking or mean score.

In our study, adequate level of knowledge was negatively associated with cataract worry and anxiety (*P* = 0.003 for both), suggesting that inadequate knowledge might be a risk factor of worry and anxiety. It is consistent with the previous study that the decision aid group had not only increased knowledge scores but alleviated anxiety for women considering birth options^41^. However, Stacey’s review showed that in many studies, there were no differences in anxiety and worry between the decision aid and control groups,^8^ implying that the increase of knowledge using decision support program would not cause undue anxiety and worry to participants.

In addition, adequate level of knowledge was closely correlated with negative attitude (*P* < 0.001) and intentions to delay cataract surgery (*P* = 0.008). It seems that this may result from increasing patients’ knowledge about the complications and risks of cataract surgery. In terms of decision quality, adequate knowledge was found negatively associated to decision conflict and inaction regret, implying that patients with increased knowledge might experience greater certainty about their options and feel more supported. It was consistent with previous studies in India^42^, Australia^43^ and the United Kingdom^41^ that knowledge improvement by decision aids helped in reducing decision conflict and uncertainty. However, there was not statistically significant in decision conflict and regret in Brazell’s study for the treatment of pelvic organ prolapse.^44^ As contradictory results of decision quality were observed including decision conflict, satisfaction, risk perception, confidence and regret in multiple decision-aid trials,^8^ further studies are needed to test out the reasons for heterogeneity of findings.

The strengths of our study lie in the detailed method about how knowledge was measured, and that was found to be responsive to psychological conditions and decision quality among cataract patients in Guangzhou. Furthermore, we used a series of validated, widely used scales to examine patients’ psychological status and their decision quality, making the results of our study more reliable.

Our study also had several limitations. First, the current study focused on people living in urban area of Guangzhou, who probably had a higher education level, greater health awareness and were more actively participated in healthcare decisions. Meanwhile, the study sample may have been biased by not recruiting those who were most fearful of surgery, less socially connected or had a problem paying for medications. As a result, the subjects included in our study may not be representative of the general population of cataract patients. Second, as cataract patients did not involve in the establishment of the knowledge scale, we may have ignored some of their demands and concerns. The third limitation of this study is its lack of intervention and follow-up investigation, so we cannot discover the cause and effect between knowledge and those factors. Further longitudinal clinical trials are needed to explore the causality and the long-term effect of decision aids on patients’ psychology and decision quality.

Knowledge level about cataract surgery is important particularly in light of new techniques such as patient decision aids to promote shared decision making. Despite this, there is limited research on how best to measure knowledge in community-based eye care screening settings. Our knowledge scale was found to be acceptable to patients with age-related cataract and responsive to a range of psychometric measurements and decision quality. Further work is needed to evaluate the validity of this knowledge scale in the context of the Chinese eye care system.

## Data Availability

All data included in this manuscript is available.

